# A pragmatic, decentralized trial of the home-based InTandem^®^ neurorehabilitation system: analyses of engagement, safety, and effectiveness from the OrcHESTRAS trial

**DOI:** 10.64898/2026.03.13.26348352

**Authors:** Louis N. Awad, Sabrina R. Taylor, Ryan T. Pohlig, Yuri A. Maricich, Seth P. Finklestein, Erika H. Riley, Cecilia A. Carlowicz, Brian A. Harris, Francois A. Bethoux

## Abstract

**Background:** Chronic stroke-related gait impairment remains a major source of disability. InTandem is an autonomous neurorehabilitation system delivering individualized, progressive rhythmic auditory stimulation for home-based gait rehabilitation.

**Objectives:** To evaluate: (1) engagement during a 12-week autonomous, home-based intervention, (2) changes in walking endurance and functional mobility, and (3) outcome differences across pre-defined engagement and baseline speed subgroups.

**Methods:** This pragmatic, decentralized trial enrolled adults ≥6 months post-stroke with residual gait deficits. Participants were asked to complete 30-minute sessions 3x/week for up to 12 weeks. Engagement was primarily assessed as the proportion achieving moderate-to-high weekly usage (> 4 weeks; benchmark p1 = 0.60). Changes in 6-Minute Walk Test (6MWT) distances and Timed Up and Go (TUG) times were analyzed using linear mixed-effects models.

**Results:** Of the 204 who initiated the intervention, 81.9% (95% CI [0.76-0.87]) engaged at least 4 weeks, meeting the primary endpoint (p < 0.001). Overall, 58.1% achieved high engagement (> 9 weeks), 23.9% moderate engagement (4-8 weeks), and 18.1% low engagement (≤3 weeks). Significant improvements in 6MWT distance (+ 26.1 ±5.6 m; 95% CI [14.99, 37.22]) and TUG times (−1.45±0.31 s; 95% CI [−2.06, −0.84]) (p < 0.001) were observed. Engagement influenced effectiveness: each additional week engaged predicted a 5.82 m greater gain in the 6MWT (SE = 2.05; 95% CI [1.77, 9.87], p < 0.005).

**Conclusions:** Autonomous home-based delivery of music-based rhythmic auditory stimulation achieved moderate-to-high engagement and improved walking endurance and functional mobility, supporting InTandem as a scalable approach to chronic stroke gait rehabilitation.

**Trial registration:** Trial registration: Clinicaltrials.gov NCT06051539. Registered on 20 September 2023. https://clinicaltrials.gov/study/NCT06051539

## 1. Introduction

Stroke is a leading cause of long-term disability worldwide,^1^ affecting millions of individuals in the United States.^2^ Persisting gait impairments lead to characteristically slow, asymmetric, and metabolically costly walking,^3–5^ and are associated with reduced community ambulation, increased fall risk, greater comorbidities, and diminished quality of life.^6–9^ Although clinical guidelines emphasize continued rehabilitation in the chronic phase,^10,11^ many survivors do not receive sufficient outpatient therapy due to barriers such as cost, transportation, and limited skilled care.^12,13^ Consequently, a critical public health priority is the development of high-efficacy interventions to improve the walking function of individuals with chronic stroke-related gait impairment. Recent expert consensus highlights an urgent need for innovative approaches that can autonomously deliver progressive, individualized, and accessible rehabilitation interventions that extend neurorehabilitation beyond the clinic into the home environment.^14^

Recent technological innovations have enabled the advancement of the InTandem neurorehabilitation system (MedRhythms, Inc.; MR-001). InTandem automates an evidence-based neurorehabilitation intervention based on auditory-motor entrainment, a neurological process in which movement non-volitionally synchronizes with rhythmic auditory cues, such as those found in music or a metronome.^15,16^ Rhythmic auditory stimulation (RAS) interventions leverage auditory-motor entrainment as a mechanism to facilitate activation and reinforcement of residual neural networks to treat gait impairments. The clinical evidence for RAS interventions includes improved walking speed and gait quality.^17–22^ Comparative studies suggest that music-based rhythmic cueing may be more effective than metronome-only cueing.^23^ Unlike metronomic cues, music engages broader neural networks beyond temporal gait regulation and may support sustained participation in rehabilitation, particularly when familiar music is used.^24–35^

Historically, the clinical impact of RAS interventions has been limited by scalability and accessibility issues. These have, in part, arisen from a dependence on in-person delivery by skilled clinicians who manually administer auditory cues based on clinical observations.^15,16^ To address this gap, InTandem integrates shoe-worn inertial sensors with decision-making algorithms that harness auditory-motor entrainment to autonomously deliver an individualized and progressive music-based RAS intervention in the home environment.^36^ During each intervention session, InTandem provides music-based RAS that adapts, in real time, to changes in each user’s entrainment and gait quality. Gait-related measurements associated with fall risk (i.e., symmetry and step-to-step variability) are continuously tracked, with musical tempo and beat salience adjusted to ensure safe intervention progression only when stable entrainment and preserved gait quality are demonstrated.^36–38^

This manuscript is the first in a series reporting the findings of the Outcomes and Health Economics of Stroke using Rhythmic Auditory Stimulation (OrcHESTRAS) study (see previously published protocol^39^). The OrcHESTRAS study builds on multiple prior studies of InTandem, including a pivotal multi-site randomized controlled trial demonstrating clinically-meaningful improvements in walking speed after five weeks of the InTandem intervention, with InTandem users 3.7x more likely than an active walking control to both exceed the 0.16 m/s minimal clinically important difference for walking speed^40^ and improve their ambulation status to “community ambulator”.^36^ Importantly, improvements in walking speed continued to accrue over the five-week intervention period without evidence of plateauing and without increased falls or adverse events (AEs) compared to the active walking control group. Biomechanical studies of the InTandem intervention complement these findings, with improvements in walking speed, gait symmetry, and the energetic cost of walking observed following a single 30-minute InTandem session.^37^ Together, these studies establish a strong mechanistic and clinical basis for additional study of the InTandem intervention. However, expanding access to home-based neurorehabilitation does not guarantee that patients will use these interventions consistently; patient engagement remains a critical determinant of real-world effectiveness.

This first manuscript from the OrcHESTRAS study builds on the foundational clinical evidence previously published to evaluate InTandem’s effects on other measurements of walking function (i.e., walking endurance and functional ability vs. walking speed), the effects of a longer intervention duration (12 weeks vs. 5 weeks), subgroup differences across a larger and broader population (n = 232 vs. n = 87), and engagement in implementation-relevant contexts (home vs. clinic). Critically, engagement is this study’s primary endpoint given that the clinical impact of home-based neurorehabilitation depends not only on therapeutic efficacy but also on if patients will use the intervention in home and community settings as intended. Prior studies of post-stroke physical activity and exercise programs have reported substantial challenges with sustained participation, with adherence rates commonly ranging from approximately 50% to 75% even in the context of structured, coached interventions.^41^ Barriers such as stroke-related impairments, fatigue, transportation limitations, and reduced motivation further constrain engagement in outpatient and home-based programs.^42^ While measures of adherence typically quantify completion of a prescribed ‘dose’, we define and examine engagement as the degree of interaction with InTandem, which is especially relevant given the unsupervised, home-based context of the intervention. Furthermore, we examine the relationship between engagement and walking outcomes to inform the broader question of how InTandem-delivered RAS could be optimally integrated into home-based chronic stroke rehabilitation. This approach is grounded in the premise that repeated, sustained, rhythmically cued gait practice in a familiar environment with familiar music may promote the neuroplasticity required to support continued functional recovery.

Our primary objective was to assess engagement with InTandem, and we hypothesized that at least 60% of participants would achieve at least moderate engagement over the 12-week intervention period. The 60% proportion threshold was selected as a pragmatic benchmark reflecting a balance between feasibility and clinical relevance. Our secondary objective was to evaluate whether the intervention improves walking endurance, as measured by the 6-Minute Walk Test (6MWT). We hypothesized that participants would demonstrate significant improvements in endurance from baseline to 12 weeks. Finally, our exploratory objectives included assessing changes in functional mobility via the Timed Up and Go (TUG) test and evaluating differential effects across subgroups. We hypothesized that clinical gains in both walking endurance and functional mobility would be enhanced among individuals with greater baseline gait impairment or those with higher levels of intervention engagement. The findings of this study will build on the established feasibility, acceptability, and effectiveness of the home-based InTandem intervention.

### 2. Methods

### 2.1 Participants

Participants were enrolled in the Outcomes and Health Economics of Stroke using Rhythmic Auditory Stimulation (Or-cHESTRAS) trial, a pragmatic, decentralized, longitudinal clinical trial evaluating engagement, clinical, and economic outcomes associated with autonomous home use of the InTandem neurorehabilitation system in individuals with chronic stroke gait impairment. The full protocol was published previously.^39^ In summary, eligible participants were ≥ 6 months post-stroke with residual gait deficits, aged 18-85 years, able to ambulate without physical assistance from another individual (assistive devices permitted), and capable of walking at least 0.4 m/s during the 6MWT. Exclusion criteria included non-reciprocal gait patterns, significant comorbid neurologic conditions affecting gait (e.g., Parkinson disease, multiple sclerosis), more than two falls in the prior month, hearing impairments precluding perception of rhythmic cues, or medical instability preventing safe participation. All participants provided electronic informed consent. The study was approved by the Advarra Institutional Review Board (Protocol #Pro00073144) and conducted in accordance with Good Clinical Practice. The InTandem system was determined to be non-significant risk under 21 CFR 812.3(m).

### 2.2 Intervention and Data Collection

Participants utilized InTandem over a 12-week period in the home environment of their choice, with a target schedule of three 30-minute sessions per week. Upon confirmation of eligibility for the study, the device was shipped to the participant’s home, and they received a guided setup call to review study expectations and how to get started with the device. Device usage data, including session timestamps and total walking minutes, were automatically uploaded to secure study servers and used to evaluate engagement (primary end-point) during Step 1 of this trial. Clinical assessments were conducted at baseline (T1) and after the 12-week intervention (T2) at designated assessment centers. Walking performance measures included walking endurance (6MWT; secondary end-point) and functional mobility (TUG; exploratory outcome). Additional patient-reported outcomes (e.g., Patient Health Questionnaire-8, Barthel Index, Patient-Reported Outcomes Measurement Information System Social Isolation Measure) and additional data collection timepoints are intended to be reported in subsequent publications. AEs were monitored throughout the course of the study and classified by severity, seriousness, and relatedness to the intervention. Falls were prospectively monitored as an adverse event of special interest during the study. Complete procedures on AE reporting for this study were described previously.^39^

### 2.3 Data Processing

Device-recorded data were exported and processed using Python (v3.11). For each participant, total weeks engaged, sessions completed, and cumulative minutes walked were computed.

- Weeks engaged: number of distinct weeks with ≥ 1 completed session starting upon device receipt.
- Sessions completed: total number of sessions during the 12-week intervention period.
- Minutes walked: cumulative minutes across all completed sessions.

Engagement was categorized as *Low, Moderate*, or *High* for each metric using predefined thresholds corresponding to one-third and two-thirds of the maximum engagement for each metric. That is, based on the guidance to complete 30-minute sessions, 3 times per week, over 12 weeks, maximum engagement would be: 12 weeks, 36 sessions, and 1080 minutes. *High* engagement would be reflected by completion of at least two thirds of this maximum engagement, *Moderate* engagement would be reflected by approximately one third to two thirds, and *Low* engagement would be reflected by completion of less than one third. More specifically, low, moderate, and high weekly engagement corresponds to ≤ 3 weeks, 4-8 weeks, and ≥ 9 weeks, respectively. Similarly, session-based engagement subgroups would be based on ≤ 11 sessions, 12-24 sessions, and ≥ 25 sessions. Finally, minute-based subgroups would be based on ≤ 359 minutes, 360-719 minutes, and ≥ 720 minutes. Participants classified as *Moderate* or *High* on a given engagement metric were considered to have achieved moderate-to-high engagement on that metric. Quality checks verified timestamps, durations, and cumulative totals; incomplete or corrupted logs were excluded, and affected participants were conservatively classified as non-engaged for the relevant metric.

### 2.4 Statistical Analysis

Analyses followed an intention-to-treat framework, with endpoint-specific analytic sets defined as:

- Engagement endpoints: all participants who initiated at least one InTandem session.
- Clinical endpoints: all participants with a baseline observation.

Missing data were assumed missing at random; outcomes were estimated via maximum likelihood. Missing baseline covariates were imputed (numeric by mean; categorical by mode).

#### Engagement Endpoints

The primary endpoint was the proportion achieving moderate-to-high engagement, assessed primarily for weeks engaged (i.e., > 4 weeks with at least one session), and secondarily for sessions completed and minutes walked. For each metric, a one-sided exact binomial test evaluated whether the observed proportion exceeded *p1 =*0.60 (success benchmark). If met, sensitivity analyses used more stringent criteria (i.e., *p1 =* 0.70, 0.80). We report the proportion, 2-sided Clopper-Pearson 95% confidence interval, and 1-sided p-value vs. *p1*.

#### Clinical Endpoints

Changes in 6MWT distance (m) and TUG time (s) from T1 to T2 were analyzed using linear mixed-effects models (random subject intercepts). Fixed effects included Time (T1, T2) and covariates Age, Sex, Time since stroke, and Assistive device use (None vs Yes). Models were fitted by maximum likelihood; Akaike Information Criterion/Bayesian Information Criterion were used to confirm parsimony. Estimated marginal means (±SE) and 95% CIs were derived from fixed effects; α = 0.05 (two-sided). Moderation of 6MWT gains was evaluated by testing Time x Engagement and Time x Speed interactions, with Engagement and Speed evaluated as continuous variables and *a priori* subgroups; results expressed as adjusted mean differences (Δ ±95% CI).

#### Engagement Subgroups

Low (< 4 weeks) vs Moderate-to-High (≥ 4 weeks) engagement.

#### Speed Subgroups

Limited (< 0.80 m/s) vs Unlimited (≥0.80 m/s) community ambulators.

## 3. Results

### 3.1 Participant baseline characteristics and InTandem intervention timeline

A total of 232 participants were eligible and enrolled; 204 participants initiated the InTandem intervention and were included in all engagement analyses. See **Table 1** for complete participant baseline characteristics. On average (±standard deviation), the 204 participants completed their virtually-guided setup of the InTandem system 2.04 ± 1.92 days after it arrived in the mail to their home and completed their first intervention session 2.86 ± 4.74 days after the device setup. During the device setup call, participants reported planning to utilize a wide range of locations for their sessions, including both outside locations such as neighborhoods/communities, parks, waterfronts, tracks, sidewalks, trails/paths and inside locations like home/kitchen/living room, mall, gym, apartments, hallways, and office/work.

**Table 1:** Participant baseline characteristics.

| Characteristic | n = 232<br>total enrolled | n = 204<br>initiated InTandem |
| --- | --- | --- |
| Age (years) | 60.8 ± 10.5 | 60.7 ± 10.7 |
| Sex |  |  |
| Male | 134 (57.8) | 115 (56.4) |
| Female | 98 (42.2) | 89 (43.6) |
| Race |  |  |
| White | 143 (61.6) | 128 (62.7) |
| Black or African American | 71 (30.6) | 61 (29.9) |
| Other / Multiple races | 18 (7.8) | 15 (7.4) |
| Ethnicity |  |  |
| Hispanic or Latino | 20 (8.6) | 17 (8.3) |
| Education (years) | 14.9 ± 2.0 | 14.9 ± 2.0 |
| Body Mass Index (kg/m<sup>2</sup>) | 30.7 ± 6.7 | 30.8 ± 6.7 |
| Time since stroke (years) | 5.6 ± 5.6 | 5.5 ± 5.2 years |
| Baseline walking speed (m/s) (derived from 6MWT) | 0.81 ± 0.30 | 0.80 ± 0.30 |
| Gait subgroup |  |  |
| Limited community ambulators (< 0.80 m/s) | 128 (55.2) | 115 (56.4) |
| Unlimited community ambulators (≥ 0.80 m/s) | 104 (44.8) | 89 (43.6) |
| Primary assistive device |  |  |
| None | 131 (56.5) | 113 (55.4) |
| Cane | 76 (32.8) | 68 (33.3) |
| Walker | 25 (10.8) | 23 (11.3) |

### 3.2 Engagement Analyses

Overall, participants accumulated 104,946 minutes of InTandem-based gait rehabilitation, engaging with the InTandem system across 4,384 unique sessions over 1,674 weeks. At the group level, participants demonstrated sustained use of InTandem, with an average (± standard deviation) of 8.2 ± 3.8 weeks engaged, 21.5 ± 15.3 sessions completed, and 514.4 ± 427.6 cumulative minutes. Participants who engaged for ≤ 3 weeks (Low, n = 37), completed an average of 1.9 ± 0.9 weeks, 2.6 ± 1.7 sessions, and 49.8 ± 44.5 minutes. Participants who engaged for 4-8 weeks (Moderate, n = 49), completed an average of 6.0 ± 1.5 weeks, 11.3 ± 4.9 sessions, and 234.1 ± 139.4 minutes. Participants who engaged for ≥ 9 weeks (High, n = 118), completed an average of 11.1 ± 1.1 weeks, 31.7 ± 11.6 sessions, and 776.5 ± 371.9 minutes.

The primary endpoint was met **(Table 2)**: 167 of 204 participants (81.9%, 95% CI [0.76, 0.87]) engaged with InTandem at least 4 weeks, exceeding p_1_ = 0.60 and p_1_ = 0.70 (p’s < 0.001), but not p_1_ = 0.80 (p = 0.29). For the session-based engagement analysis, 138 of 204 participants (67.6%, 95% CI [0.61, 0.74]) engaged with InTandem at least 12 sessions, exceeding p_1_ = 0.60 (p = 0.015) but not p_1_ = 0.70 (p = 0.254). Finally, for the minute-based engagement analysis, 113 of 204 participants (55.4%, 95% CI [0.48, 0.62]) engaged with InTandem at least 360 minutes, with the proportion not exceeding p_1_ = 0.60 (p = 0.102).

**Table 2:**
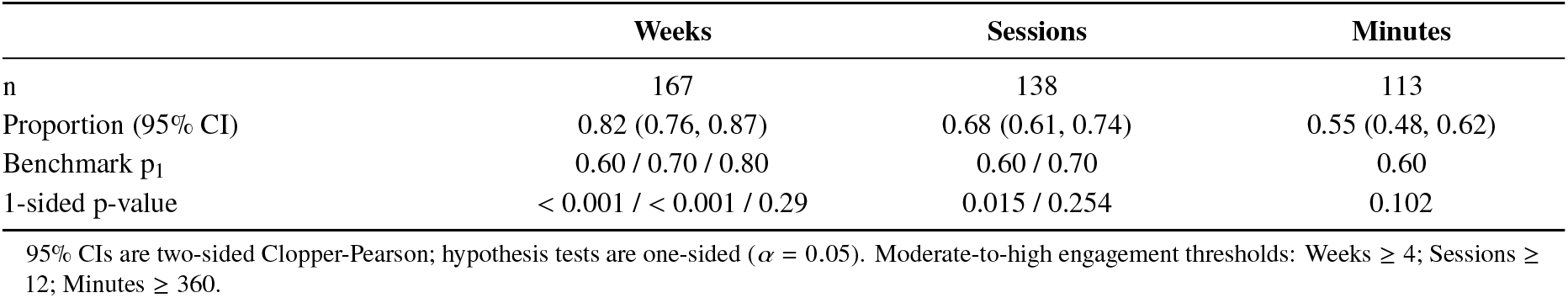
Proportion of participants achieving moderate-to-high engagement (n = 204)

|  | Weeks | Sessions | Minutes |
| --- | --- | --- | --- |
| n | 167 | 138 | 113 |
| Proportion (95% CI) | 0.82 (0.76, 0.87) | 0.68 (0.61, 0.74) | 0.55 (0.48, 0.62) |
| Benchmark p<sub>1</sub> | 0.60 / 0.70 / 0.80 | 0.60 / 0.70 | 0.60 |
| 1-sided p-value | < 0.001 / < 0.001 / 0.29 | 0.015 / 0.254 | 0.102 |

95% CIs are two-sided Clopper-Pearson; hypothesis tests are one-sided ( $\alpha = 0.05$ ). Moderate-to-high engagement thresholds: Weeks $\geq 4$ ; Sessions $\geq 12$ ; Minutes $\geq 360$ .

### 3.3 Secondary Clinical Endpoint and Subgroup Analyses: 6-Minute Walk Test Distance

Linear mixed-effects models for the 6MWT included all enrolled participants regardless of completion (N = 232 participants; 391 observations) and adjusted for Age, Sex, Time since stroke, and Assistive device use. The adjusted T1 to T2 change in the 6MWT was +26.1 ± 5.6 m (95% CI [14.99,37.22], p < 0.001).

#### Engagement moderation analyses

Engagement influenced improvements in 6MWT distance (Time × Engagement: F(1,167.18) = 8.06, p = 0.005). The interaction estimate indicated a dose-response relationship, such that each additional week of engagement was associated with +5.82 m *greater* improvement in 6MWT distance from T1 to T2 (β = 5.82 m/week, SE = 2.05; 95% CI [1.77, 9.87]; **Figure 1A**). When engagement was modeled using *a priori* sub-groups (≥ 4 weeks vs < 4 weeks), the Time × Engagement Group interaction was statistically and clinically meaningful (F(1,177.5) = 4.22, p = 0.041). In brief, the Moderate-to-High Engagement subgroup, which had an average 9.6 ± 2.6 weeks engaged, showed a significant improvement of +31.6 ± 5.9 m (F(1,149.3) = 29.12, p < 0.001). In contrast, the Low Engagement subgroup, which had a modest 1.9 ± 0.9 weeks engaged, showed a non-significant decline of −27. 0 ± 27.9 m (F(1,178.8) = 0.94, p = 0.335) **(Figure 1B)**. The between-group difference of approximately + 58.6 m exceeds the 34.4 m minimal clinically important difference.^43^

**Figure 1.**
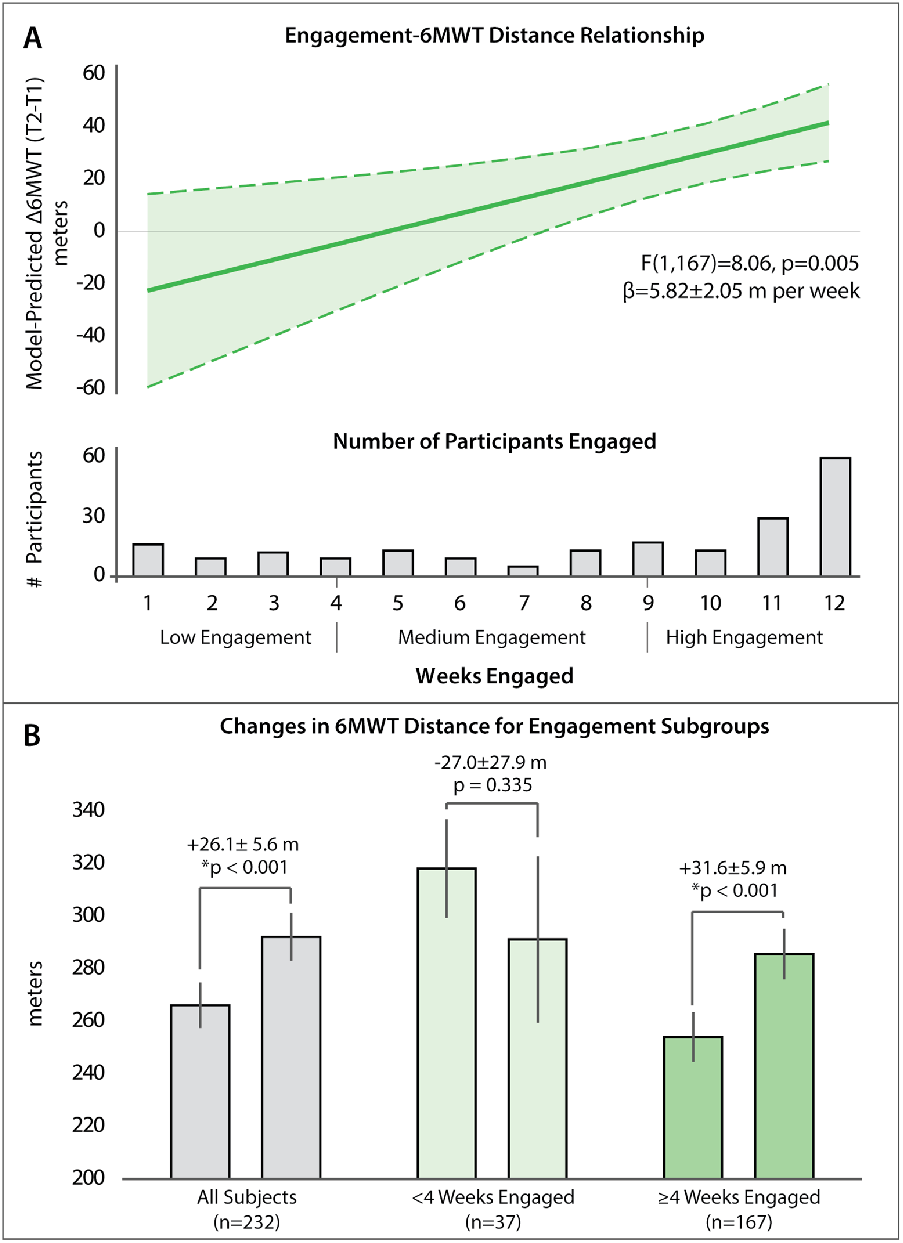
(A) Model-predicted change in 6MWT distance (Δ6MWT = T2 - T1) according to weeks engaged. The solid line indicates the estimated marginal effect, and dashed lines with the shaded region indicate 95% confidence intervals, from a linear mixed-effects model adjusted for age, sex, assistive device category, and time since stroke. The histogram below displays the number of participants contributing data at each engagement duration. (B) Model-adjusted pre- and post-intervention 6MWT distances in the full cohort and prespecified engagement subgroups. Bars show adjusted mean values (m), error bars represent standard errors, and brackets indicate adjusted within-group change.

#### Speed moderation analyses

Baseline walking speed did not significantly moderate improvements in 6MWT distance over the intervention period (Time × Speed: F(1,289.95) = 2.12, p = 0.146). Consistent with this finding, the subgroup analysis showed a non-significant Time × Speed Category (< 0.8 m/s vs ≥ 0.8 m/s) interaction (F(1,159.77) = 3.24, p = 0.074). However, examination of 6MWT changes within each subgroup revealed that the Limited community ambulator subgroup (< 0.8 m/s) demonstrated a significant improvement of +34.9 ± 7.4 m (F(1,155.23) = 22.55, p <.001), whereas the Unlimited community ambulator subgroup (≥0.8 m/s) showed no significant change (+14.3 ± 8.7 m; F(1,163.00) = 2.70, p = 0.102).

### 3.4 Exploratory Clinical Endpoint and Subgroup Analyses: Timed Up and Go Time

Linear mixed-effects model for TUG time (N = 232 participants; 390 observations) similarly revealed a significant improvement after adjusting for Age, Sex, Time since stroke, and Assistive device use. The adjusted T1 to T2 change in TUG time was −1.45 ± 0.31 s (95% CI [−2.06, −0.84], p <0.001).

#### Engagement moderation analyses

Engagement did not significantly moderate TUG improvements. When modeled as a continuous variable, the Time × Engagement interaction was not significant (F(1,158.5) = 0.15, p = 0.696), with a near-zero interaction estimate (β = −0.05 ± 0.11 s/week). Consistent with this, the subgroup model (≥ 4 weeks vs < 4 weeks) showed no evidence of significant moderation (Time × Engagement: F(1,165.5) = 0.20, p = 0.655). However, within-group analyses showed that the Moderate-to-High Engagement subgroup (≥ 4 weeks) demonstrated a significant improvement of −1.39 ± 0.33 s (F(1,138.3) = 18.37, p < 0.001), whereas the Low Engagement subgroup (< 4 weeks) showed a non-significant improvement of −2.10 ± 1.54 s (F(1,167.8) = 1.85, p = 0.175).

#### Speed moderation analyses

Baseline walking speed did not significantly moderate TUG improvements. When baseline walking speed was modeled as a continuous variable, the Time × Speed interaction was not significant (F(1,155.9) = 0.86, p = 0.355). The subgroup analysis (< 0.8 m/s vs ≥ 0.8 m/s) was consistent, also showing no significant moderation (Time × Speed Category: F(1,150.5) = 3.14, p = 0.079). However, within-group analyses showed that the Limited community ambulator subgroup (< 0.8 m/s) significantly improved by −1.92 ± 0.40 s (F(1,147.6) = 22.87, p < 0.001), whereas the Unlimited community ambulator subgroup (≥ 0.8 m/s) showed a smaller, non-significant improvement of −0.82 ± 0.48 s (F(1,152.5) = 2.94, p = 0.088).

### 3.5 Adverse Events

Adverse events were tracked prior to device use. The time between the day of enrollment to the day of first device use was, on average, 3.8 ± 1.2 weeks. None of the AEs and serious AEs reported during this time were considered related to the intervention. Forty-one AEs were reported during this time period; three were serious AEs, which were classified as severe. Six falls of mild to moderate severity were also reported during this period. The severe serious AEs and falls captured during this pre-intervention phase were consistent with the baseline medical and fall risk of individuals with chronic stroke-related gait impairment.

During the 12-week intervention period, a total of 137 AEs were reported, of which 17 (12.4%) were classified as serious AEs. All serious AEs were reported as severe; 15 (88.2%) were assessed as unrelated to the intervention, and two were considered possibly related. The two possibly related AEs were seizures occurring in a single participant with a known history of epilepsy and temporally associated with device use. No deaths or life-threatening events occurred during this 12-week intervention phase.

Twenty-seven falls were also reported during the intervention phase, the majority of which were mild or moderate in severity and assessed as unrelated or unlikely to be related to the device (85.2%). Four falls (14.8%) were classified as possibly (n = 3) or definitely (n = 1) related to device use. The fall definitely related to device use occurred during a walking session, resulting in skin abrasions and a fracture, and subsequent study withdrawal. Across all phases reported here, 28 participants reported at least 1 fall (13.7%), and three participants (1.5%) reported more than 1 fall. No additional fall-related serious AEs were observed.

## 4. Discussion

This study demonstrates that sustained engagement with an autonomous, home-based RAS intervention is achievable in adults with chronic stroke and is associated with improved walking endurance and functional mobility. In this cohort of 204 participants, 82% achieved at least four weeks of engagement, exceeding our 60% feasibility benchmark. These findings support InTandem as a scalable approach to home-based gait rehabilitation.

### Transforming the neuroscience of music to a scalable and accessible intervention

RAS and music-based gait interventions have a strong scientific foundation, with more than three decades of evidence demonstrating improvements in walking speed, temporal and spatial symmetry, and coordination.^15–22,34^ Expert consensus and guideline statements increasingly recognize RAS as an evidence-based option for gait rehabilitation.^10,11,14^ Despite this, RAS remains underutilized and inaccessible in routine stroke care, largely because traditional delivery models rely on skilled clinicians providing rhythmic cues during in-person sessions. InTandem represents a technological advancement that addresses this access gap; the technology delivers RAS autonomously through a sensor-driven, closed-loop system operated in the home environment without the need for clinician oversight. The trial shows that when placed directly in the hands of stroke survivors, most participants engage with the intervention consistently enough to achieve meaningful improvements in walking function, with a safety profile consistent with expectations for this population. Together, the preponderance of evidence demonstrates that RAS is an effective intervention and InTandem is an effective, safe vehicle for delivering that intervention at scale in the home environment.

### Safety of the autonomous, home-based InTandem intervention

Safety was a critical consideration in the design of this intervention given the elevated baseline risk of adverse events and falls among individuals with chronic stroke. Across the 12-week intervention period, most AEs were assessed as unrelated or unlikely to be related to device use and were consistent with the background rate of medical events observed prior to intervention initiation. Serious AEs were infrequent and predominantly unrelated to the intervention. Two serious adverse events classified as possibly related to the intervention were seizures occurring in a single participant with a known history of epilepsy. Although causality cannot be established, they were temporally associated with device use and seizures are a known risk in individuals with chronic stroke and pre-existing epilepsy, and were thus monitored carefully throughout the study.

Falls were observed before and during the intervention period, with most falls during the intervention period mild and not attributable to device use. Fall incidence (< 14%) and recurrent falls (< 2%) were lower than rates commonly reported among community-dwelling individuals with chronic stroke, where 23-46% of individuals fall at least once annually and recurrence is substantially more common than in the general older adult population.^44–47^

Importantly, these findings were observed across more than 100,000+ minutes of autonomously delivered gait rehabilitation in a large, geographically distributed cohort. Together, these results support the tolerability of extended home-based RAS intervention when used in individuals meeting the device’s intended-use criteria and following the instructions for use.

### Engagement with the autonomous, home-based InTandem intervention

Engagement was evaluated across minutes, sessions, and weeks. Whereas week- and session-based metrics capture repeated return to the intervention, minutes-based engagement may be more sensitive to fatigability, endurance limitations, or within-session motivation. The high proportion of participants achieving moderate-to-high week- and session-based engagement is notable given the well-documented challenges of sustaining rehabilitation participation in the chronic stroke phase^41,42^, emphasizing InTandem’s ability to support repeated therapeutic exposure in home-based contexts. Importantly, though there was variability in engagement, most participants were able to integrate InTandem into their home routines at levels associated with meaningful improvements in walking endurance and functional mobility.

### New evidence of clinical effectiveness complements prior safety, efficacy, and recovery rate evidence

The 12-week, home-based autonomous intervention produced clinically meaningful improvements in walking endurance and functional mobility. The observed relationship between engagement and changes in 6MWT distance suggest a dose-response improvement in walking endurance, with each additional week of engagement corresponding to an additional ~6 m improvement in 6MWT distance. When extrapolated across the 12-week intervention period, this dose-response relationship predicts the emergence of clinically meaningful improvements in 6MWT distance (i.e., > 34.4 m MCID) by approximately the 8th week of InTandem use (i.e., upper bound of 95% CI surpasses the MCID), with a mean predicted change of over 41 m by 12 weeks. The study’s finding of an engagement effect across the continuous and categorical engagement models, and the time-course-of-change analysis reported in the prior randomized controlled trial^36^, together support the interpretation that sustained engagement is a meaningful prognostic indicator of InTandem’s rehabilitative benefits.

These findings extend prior mechanistic and randomized controlled trial evidence for the InTandem intervention. A previous biomechanical study showed that a single 30-minute InTandem session can improve walking speed, temporal symmetry, and energetic cost of walking.^37^ A subsequent multi-site randomized controlled trial demonstrated that a 5-week InTandem intervention more than doubled walking speed gains compared with an active walking control, with a time course of change analysis indicating an ~0.05 m/s increase in walking speed for every six InTandem sessions.^36^ The present study complements those findings by demonstrating improved endurance and functional mobility during extended autonomous home use.

Improvements of this magnitude meet or exceed commonly reported minimal clinically important differences for the 6MWT in people with chronic stroke impairments.^43^ In practical terms, gains of 20-50 m may translate into improved community ambulation capacity, including the ability to traverse longer household and community distances such as parking lots, sidewalks, and grocery store aisles.^48^ Improved walking endurance may also reduce mobility-related participation restrictions and increase overall physical activity exposure.^49^ Importantly, interventions that meaningfully increase community mobility post-stroke may help mitigate downstream healthcare utilization associated with falls, deconditioning, and mobility-related functional decline. Autonomous home-based interventions may further enhance impact by expanding access and supporting participation in rehabilitation.

### Who benefits most? Role of baseline gait impairment

Importantly, InTandem’s intervention effects do not appear to be strongly moderated by baseline walking speed; both slower and faster individuals benefited from the intervention. Nonetheless, it is important to note that despite the between-group difference not being significant, within-group analyses showed significant and clinically-meaningful^50^ improvement (+34.9 ± 7.4 m, p <.001) only in the limited community ambulator subgroup (< 0.8 m/s). In contrast, while the unlimited community ambulator subgroup (≥0.8 m/s) also improved, the magnitude was half as small and was not significant or clinically-meaningful (+14.3 ± 8.7 m, p = 0.102). One potential explanation is that individuals with greater baseline gait impairment (captured in slower speeds) may have more capacity for improvement.^51^

### Limitations and future directions

Several limitations warrant consideration. First, this phase of the trial utilized a single-arm design without a concurrent control group. This design was selected purposefully as this study builds on a randomized controlled trial that previously established the efficacy and safety of InTandem compared with a treatment-matched active walking control group.^39^ Nevertheless, improvements in 6MWT and TUG could reflect, in part, regression to the mean, Hawthorne effects, or non-specific benefits of increased walking. However, participants were, on average, more than six years post-stroke, a time window in which spontaneous functional gains are unlikely.^13^ Furthermore, the low-engagement subgroup exhibited a decline in 6MWT distance, arguing against a uniformly positive temporal trend.

Second, engagement was categorized using pragmatic thresholds tied to fractions of the maximum recommended engagement and did not account for other potentially important characteristics of intervention exposure, like walking intensity or environmental complexity. Participants used the system in real-world settings where terrain, weather, and other environmental factors may influence walking performance. Future work could incorporate sensor-derived metrics (e.g., step counts or cadence patterns) and contextual information to better characterize engagement and its relationship to clinical outcomes.

Third, although the regression models were adjusted for key covariates, residual confounding is possible. Participants who engaged more with InTandem may also have differed in baseline motivation, activity levels, or concurrent therapies. In addition, reasons for withdrawal or loss to follow-up were not systematically captured for all participants, limiting the ability to fully interpret participant retention. Finally, this paper describes analyses from the first part of Step 1 of the OrcHESTRAS trial;^39^ additional planned analyses will examine the durability of InTandem’s intervention effect, its cost-effectiveness, and withdrawal and re-treatment effects.

## 5. Conclusions

In this large, decentralized clinical trial, the 12-week, fully autonomous home-based InTandem intervention demonstrated high-to-moderate engagement and clinically meaningful improvements in functional walking ability in adults with chronic stroke gait impairment. When combined with prior mechanistic and randomized controlled trials, these findings support InTandem as a safe, scalable, evidence-based approach to delivering RAS-based gait rehabilitation in the home environment. By transforming a well-established but traditionally unscalable neurorehabilitation intervention - RAS - into a widely accessible, home-delivered intervention, InTandem offers a pathway toward a new standard of care for chronic stroke rehabilitation that addresses gaps in both access to evidence-based rehabilitation and engagement with home-based interventions.

## Data Availability

All data produced in the present study are available upon reasonable request to the authors

## Acknowledgments

The authors extend their sincere gratitude to all the participants and their families for their time, commitment, and trust in this study. We also wish to thank our colleagues at MedRhythms, Inc. for their ongoing support, collaboration, and dedication throughout the trial. We acknowledge the valuable contributions of sub-investigators Drs. Vikram Garg and Judith Weisfuse for their clinical expertise and commitment to participant care. Finally, we acknowledge the team at Curavit Clinical Research for their expertise and operational support as our Contract Research Organization.

## Author contributions

ST, CC, BH, LA, YM, SF, ER, and FB contributed to conceptualization and study design. LA, RP, and ST contributed to data curation, formal analysis, and methodology. ST, CC, BH, LA, YM, SF, ER, and FB contributed to investigation, methodology, and supervision. ST and FB contributed to project administration, and CC contributed resources. ST and BH contributed to funding acquisition. ST contributed to validation. LA and ST contributed to writing the original draft. LA, RP, ST, YM, SF, ER, CC, BH, and FB contributed to critical review and editing of the manuscript. All authors read and approved the final manuscript.

## Statements and declarations

### Ethical considerations

This study received ethical approval from the Advarra Institutional Review (Pro00073144) on August 15, 2023.

### Consent to participate

All participants provided written informed consent prior to participating.

### Consent for publication

Not applicable

### Data availability

The datasets generated during and/or analyzed during the current study are available from the corresponding author on request.

